# Unused Samples from Clinical Blood Draws as a Resource for Maximizing Research Samples while Mitigating Iatrogenic Anemia Risks: A Pilot Study

**DOI:** 10.1101/2025.04.17.25326023

**Authors:** Ian S. Jaffe, Imad Aljabban, Jacqueline I. Kim, Nicolas Dundas, Karen Khalil, Stefany Rosa, Zasha Zayas, McKenna Nally, Estefania Gallego, Adam Griesemer, Robert A. Montgomery, Jeffrey M. Stern

**Affiliations:** Department of Surgery, New York University Grossman School of Medicine, 317 East 34th Street, New York, NY, USA 10016; Department of Surgery, Columbia University School of Medicine, 622 W 168th St, New York, NY USA 10032; Transplant Institute, New York University Grossman School of Medicine, 317 East 34th Street, New York, NY, USA 10016; Department of Pathology, New York University Grossman School of Medicine, 317 East 34th Street, New York, NY, USA 10016; Center for Biospecimen Research & Development, New York University Grossman School of Medicine, 317 East 34th Street, New York, NY, USA 10016

**Keywords:** iatrogenic anemia, xenotransplantation, biorepository, blood samples, laboratory

## Abstract

**Background:** Translational research driven by large-scale biological testing requires significant volumes of blood for research testing. However, blood is also required for clinical management of research subjects, which must take priority. Paradoxically, much of the blood drawn for clinical management goes unused. Here, we present our approach for retrieving unused blood samples collected for clinical management and recycling them for research purposes.

**Methods:** Clinical Blood samples were collected for 60 days during a 61-day xenotransplantation experiment in a brain-dead decedent. Twice weekly, research staff went to the chemistry and hematology laboratories and collected stored blood, serum, and plasma samples that were >12 hours old. Sample collection and storage before retrieval was per standard clinical protocols. Samples were de-identified and relabeled and brought to a central biorepository for processing and storage. The quantity of plasma, serum, red blood cells (RBCs), and peripheral blood mononuclear cells (PBMCs) collected from clinical labs and bespoke research blood draws were compared.

**Results:** Unused blood from clinical samples yielded a minimum of 6.0 ml per day of plasma, representing 62% of all plasma obtained. Serum was only recoverable on 13 days (22%), with a mean 2.3 ml collected on those days, representing 8% of all serum obtained. PBMCs were only recoverable on six days (10%).

**Conclusions:** Overdrawn clinical laboratory samples represent an untapped resource of blood samples for research and can help augment samples collected explicitly for research purposes. With careful planning, this represents an opportunity to minimize iatrogenic blood loss in clinical-translational research.

## Introduction

The semi-fictional scholar Heinrich Faust succinctly describes in Goethe’s *Faust, A Tragedy*: “Blut ist ein ganz besondrer Saft / *Blood is a very special juice*.”(1) Despite this truism, it has only been in recent decades that the *specialness* of blood has truly been unlocked for clinical and translational research. We can now use blood as the powerful data source, synthesizing a multitude of biochemical tests to create understanding of basic biology and develop drugs and treatments.(2) Importantly, each test run may require its own blood aliquot or even its own independent blood draw, leaving us with a simple problem: some translational and early phase clinical research—especially studies with frequent monitoring—can require a significant amount of blood.(3)

Importantly, repeated blood draws are not benign; iatrogenic anemia from diagnostic blood loss is cumulative and common, with associated clinical risks from both the anemia and blood transfusions, when required.(4–6) When frequent blood testing is required for both research and clinical purposes, the amount of blood drawn must be limited to mitigate some of these risks. Ethically, clinical management must be prioritized over research. Yet, the reality is that the majority of blood drawn for clinical testing—often as much as 4 ml per tube—is unnecessary and ends up being discarded.(7) In fact, the volume of blood drawn for many clinical tests is an order of magnitude greater than the volume required.(8) While there have been numerous efforts to ameliorate these issues, such as point-of-care testing, the use of smaller volume tubes (“pediatric tubes”), or streamlined clinical decision-making to avoid duplication, clinical practice routinely still over-draws blood.(9–12) What if the overdrawn blood could instead be repurposed for research applications in subjects enrolled in clinical trials? Our research focuses on advancing xenotransplantation, which currently involves studies in brain-dead decedents. While in decedents we can accept risks of transfusions that might otherwise be unacceptable for living subjects with drawing large volumes of blood and transfusing blood back—allowing for us to fulfill all our research needs—the goal is to have xenotransplantation enter clinical trials in living recipients (which has recently begun with several expanded-access single-patient approvals),(13,14) where the aforementioned concerns limit blood draws. Thus, we aimed to use our decedent experiments to pilot the collection of unused blood from clinical testing draws, for potential future application to living recipients. Here, we will specifically review our experience collecting unused blood over the course of a two-month decedent experiment.

## Materials and Methods

### Subject Enrollment and Ethical Considerations

A brain-dead human decedent was used for this study and has been extensively previously described.(15–19) Autonomous informed consent free from coercion for whole body donation was obtained from the decedent’s next-of-kin. The decedent was kept on somatic support for a 61-day xenotransplant experiment. All research activities were conducted in accordance with guidelines set by the New York University Research on Decedents Oversight Committee and under ethical approval from that same committee.

### Collection and Processing of Blood Derivatives

Clinical blood draws were performed using standard techniques at a frequency determined by the intensive care unit clinicians. Research blood draws were performed once daily using standard techniques. Research blood was brought directly to our institution’s research biorepository for immediate processing and frozen storage. Clinical blood was brought to the hematology or chemistry laboratories per usual standards. After processing and testing was performed by these laboratories, unused blood was stored at 4°C. Twice weekly, members of the research team would retrieve blood samples that had been drawn 12-96 hours prior and transferred these samples to our institution’s research biorepository for processing and frozen storage. Samples less than 12 hours old were retained in the hematology or chemistry laboratories in case add-on tests were needed. Depending on the additives present in the blood collection tube, blood was spun and stored as serum or plasma using standard protocols. Spun red blood cells (RBCs) were also collected and stored. When feasible, peripheral blood mononuclear cells (PBMCs) were also collected and stored.

### Analysis

We limited our analysis to the first 60 days of the 61-day experiment, since additional blood samples collected on the final day of the experiment were not representative of usual practices, given that somatic support was withdrawn on that day. The volumes of plasma, serum, and spun RBCs, and the number of PBMCs are reported descriptively. We also report limited clinical and operational details. Research data are available upon request.

## Results

### Plasma and Serum

The primary objective was to obtain plasma and serum for potential biochemical testing applications. Plasma was able to be recovered from clinical labs every day of the experiment, with a minimum of 6.0 ml per day (Figure 1A). This represented 62% of all plasma obtained during the experiment. Serum, meanwhile, was only recoverable on 13 days (22%), with a median of 2.5 ml (IQR 1.5-3.0 ml) collected on days where serum was recoverable (Figure 1B). This represented 8% of all serum obtained during the experiment.

**Figure 1.**
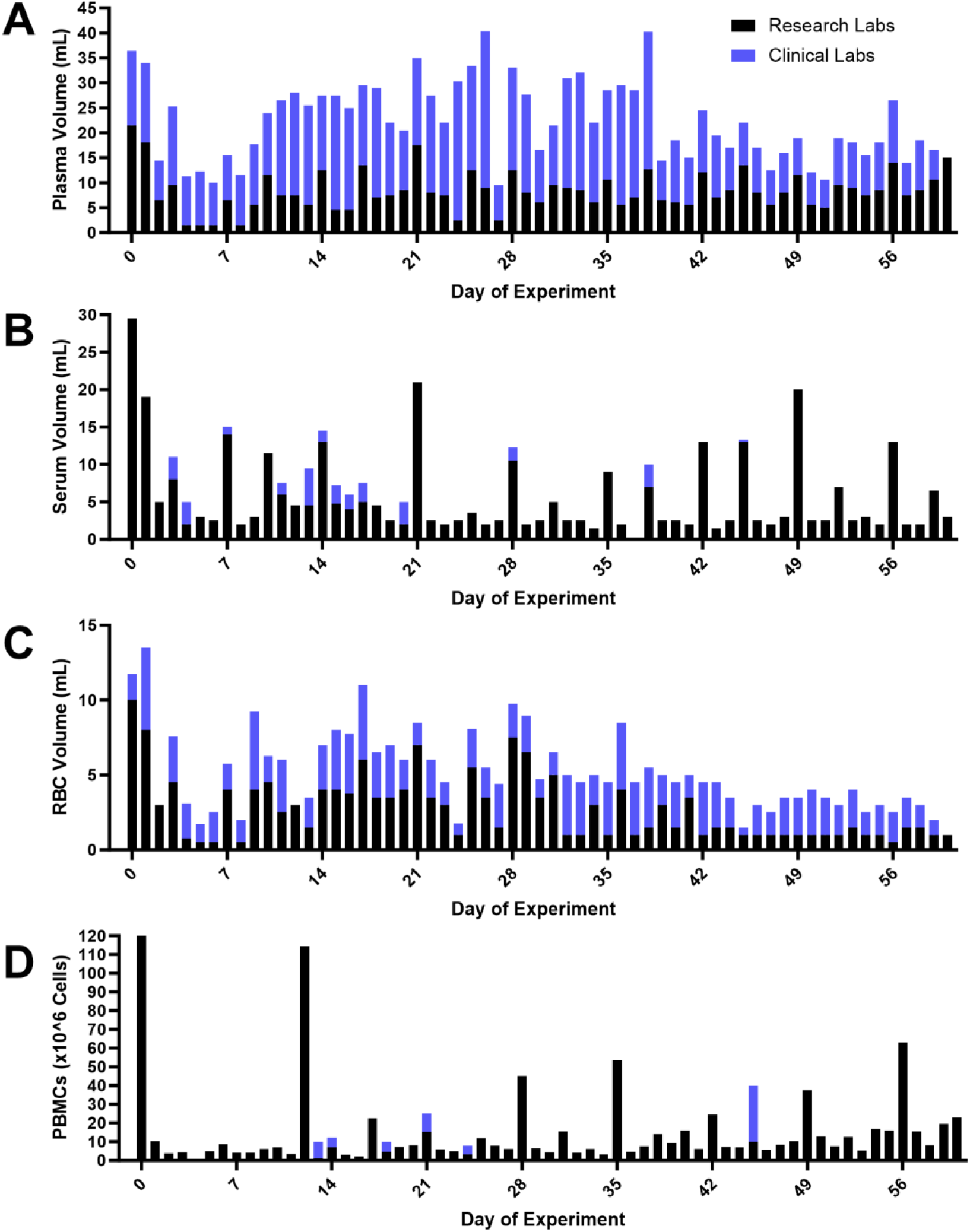
Quantity of blood derivatives obtained for research from clinical and research lab draws. Plasma, serum, red blood cells (RBCs), and peripheral blood mononuclear cells (PBMCs) were isolated from blood obtained explicitly for research (“Research Labs”) and blood initially drawn for clinical management that after going unused was retrieved by the research team (“Clinical Labs”). The quantity of A) Plasma, B) Serum, C) RBCs, and D) PBMCs collected from each day of the 60-day experiment is shown.

### Red Blood Cells

Although mainly a by-product of spinning down blood to obtain plasma and serum, spun red blood cells (RBCs) have some research applications. RBCs were obtained on 58 days of the experiment (97%), with a median of 2 ml (IQR 1.5-3.05 ml) of RBCs recovered per day (Figure 1C). This represented 47% of the volume of spun RBCs obtained for research purposes during the experiment.

### Peripheral Blood Mononuclear Cells

We sought to obtain peripheral blood mononuclear cells (PBMCs) for several immunology research applications. Since PBMCs cannot be obtained from already-spun tubes, PBMCs were rarely recoverable from samples processed by the clinical laboratories. There were only six individual days (10%) on which a single tube of blood was available for PBMC recovery (Figure 1D). A mean 10.6 x10^6 PBMCs were obtained per tube, although this was highly variable in the context of multimodal immunosuppression.

### Blood Replenishment

During the 60 days analyzed here, a total of 25 units of packed red blood cells (approximately 250 ml/unit) were required to be transfused. This was necessitated by multiple factors: anemia of chronic disease, which was present prior to the study; bone marrow suppression secondary to the immunosuppression regimen; and a deliberate decision in the study not to administer exogenous erythropoietin to address very low erythropoietin levels, aiming instead to evaluate the xenograft’s capacity to produce erythropoietin (which was not detected). Additionally, frequent blood draws for both clinical management and research purposes contributed to iatrogenic anemia. Notably, research blood draws exceeded standard clinical care requirements and were not restricted by anemia severity; instead, transfusions were used liberally to address any resulting anemia. *Operational Requirements* Approximately 26 person-hours were required to retrieve blood tubes over the 60 days. During the same period, approximately 59 person-hours were required for blood processing, aliquoting, and labeling of stored samples.

## Discussion

In this pilot study, we have demonstrated that recovering unused blood from clinical lab draws can reliably provide plasma and RBCs for research. Plasma recovery was particularly robust, often surpassing the dedicated research samples we drew. Given the fact that plasma from clinical labs was recovered at far more timepoints (average 3.3 times/day) versus one time per day for research labs, unused plasma recovered from clinical labs represented a potentially more reliable and granular source of plasma for research studies. However, other blood derivatives (in particular, serum and PBMCs) could not be reliably recovered from unused clinical blood.

The consistent recovery of plasma is unsurprising, as many clinical laboratories, including our own, utilize plasma for many common clinical tests. This preference stems from its easier technical implementation: plasma does not require waiting for blood to clot, avoids complications associated with anticoagulant medications, eliminates potential assay interference from fibrin formation, and yields a greater volume per unit of blood drawn compared to serum (20). These same advantages, which have driven its adoption in clinical practice, also make plasma an appropriate substrate for a wide range of research applications, particularly in viral detection, biomarker, metabolomic, and proteomic studies.

However, certain applications critical to transplant and xenotransplant research—notably serologies and antibody crossmatches—necessitate the use of serum tubes due to the potential interference from circulating clotting factors in plasma. Unfortunately, the inconsistent recovery of serum samples observed in this study suggests that relying on recovered serum to replace or supplement planned research draws is not feasible. There are several limitations of this study. First, this was a single-subject pilot study focused on a decedent subject undergoing intensive care and somatic support in which clinical blood draws were frequent. Importantly, as this study was conducted in a brain-dead decedent, the team was not concerned about causing iatrogenic anemia from large-volume blood draws. The decedent received 25 blood transfusions during the study, a practice not applicable to the majority of intensive care patients, given widespread patient blood management practices. The volume and frequency of blood draws observed in this study are clearly above typical levels for intensive care patients. For comparison, the average blood draw per day in intensive care patients is approximately 28.3 mL,(21) while the leftover serum and plasma recovered in our study averaged 14.4 mL. Extrapolating this to whole blood suggests an excess blood draw volume similar to the average daily blood draw in analogous patients. Although we were unable to formally evaluate the initial volume of blood drawn, it is evident that the volumes were well above typical levels. Therefore, the volume of recovered excess blood samples is almost certainly an overestimate compared to what may be seen in living patients. Nonetheless, the volumes recovered in this study likely far exceed the needs of most research applications. Second, local practices regarding the use of serum versus plasma draws and tube storage may vary significantly between hospital systems. These institutional differences could affect the feasibility and utility of residual sample collection in other settings.

Third, the quality of blood derivatives obtained through this process may be degraded by repeated handling and prolonged non-frozen cold storage. Although a recent study including some members of our group demonstrated that such samples can reliably be used for genomic DNA testing, there have been no such studies establishing the reliability of other biochemical tests on samples handled in this fashion.(22) Prior evaluations have demonstrated greater than 90% stability of the proteome when centrifuge-to-freezing times of plasma samples have been as long as 20 hours (23); however, studies evaluating the longer storage times used by our group have not been conducted. It is reasonable to believe that longer collection-to-recovery times may be detrimental for some applications, requiring further platform-specific analysis. For viral testing—another area of interest to our group—fidelity of viral DNA has been demonstrated for samples stored for up to one week, even at warm temperatures.(24,25) At the same time, the two-month sample size and consistency of blood sampling methods gives our study considerable internal validity with great extrapolation potential to other subjects and scenarios.

Unused, overdrawn blood from clinical laboratory testing represents a potentially valuable source of blood in research studies where repeated monitoring or chronic anemia must limit the volume of blood that can be drawn. We have shown that plasma from such sources can potentially supplant the need to draw additional plasma for research, allowing for iatrogenic harm to be partially mitigated. In future studies, we plan to validate that plasma recovered from clinical laboratories is reliable for biochemical testing. Future clinical trials could incorporate strategies for recovering unused blood from clinical draws, prioritizing research utility while actively reducing iatrogenic harm. Validating the reliability of these samples for various biochemical tests and multiomics analyses will be a crucial next step in ensuring their applicability to diverse research scenarios.

## Author Contributions

Conceptualization – ISJ, IA, JIK, KK, JMS. Investigation – ISJ, IA, JIK, ND, KK, ZZ, MN, EG. Resources – ISJ, KK, SR, ZZ, MN, AG, RAM. Data Curation – ISJ, ND. Writing – Original Draft – ISJ, JIK, KK, JMS. Writing – Review & Editing – ISJ, IA, JIK, ND, KK, SR, ZZ, MN, EG, AG, JMS. Visualization – ISJ. Supervision - KK, AG, RAM, JMS. Project Administration – ISJ, IA, JIK, ND, KK, JMS. Funding Acquisition - AG, RAM.

## Acknowledgments

The authors wish to thank the staff of the Center for Biospecimen Research and Development (CBRD) and NYU Langone Health for their tireless support of the parent project this manuscript is based on, including Britney A. Paredes Lopez, Jonathan Kim, Sandra Mendoza, Bernice Pham, and Andre Moreira.

## Data Availability Statement

Data are available upon request to the corresponding author.

## References

1. Von Goethe JW. Faust: eine tragödie. Vol. 1. T. Stroefer; 1879.

2. Subramanian I, Verma S, Kumar S, Jere A, Anamika K. Multi-omics data integration, interpretation, and its application. Bioinformatics and biology insights. 2020;14:1177932219899051.

3. Poitout-Belissent F, Aulbach A, Tripathi N, Ramaiah L. Reducing blood volume requirements for clinical pathology testing in toxicologic studies—points to consider. Veterinary Clinical Pathology. 2016;45(4):534–51.

4. Wisser D, van Ackern K, Knoll E, Wisser H, Bertsch T. Blood loss from laboratory tests. Clinical chemistry. 2003;49(10):1651–5.

5. Helmer P, Hottenrott S, Steinisch A, Röder D, Schubert J, Steigerwald U, et al. Avoidable Blood Loss in Critical Care and Patient Blood Management: Scoping Review of Diagnostic Blood Loss. Journal of Clinical Medicine. 2022 Jan;11(2):320.

6. Hébert PC, McDonald BJ, Tinmouth A. Clinical consequences of anemia and red cell transfusion in the critically ill. Critical Care Clinics. 2004 Apr 1;20(2):225–35.

7. Hicks JM. Excessive blood drawing for laboratory tests. New England Journal of Medicine. 1999;340(21):1690–1690.

8. Dale JC, Ruby SG. Specimen collection volumes for laboratory tests: A College of American Pathologists study for 140 laboratories. Archives of Pathology & Laboratory Medicine. 2003 Feb;127(2):162–8.

9. Geaghan S. Diagnostic Test Sample Volume: How Much Is Too Much?: Leveraging Point-of-Care Testing to Reduce Sample Volumes, Iatrogenic Anemia, and Transfusion Requirement in Adult Populations. Point of Care. 2011;10(4):157–62.

10. Myles N, von Wielligh J, Kyriacou M, Ventrice T, To LB. A cohort study assessing the impact of small volume blood tubes on diagnostic test quality and iatrogenic blood loss in a cohort of adult haematology patients. Internal Medicine Journal. 2018;48(7):817–21.

11. Whitehead NS, Williams LO, Meleth S, Kennedy SM, Ubaka-Blackmoore N, Geaghan SM, et al. Interventions to prevent iatrogenic anemia: a Laboratory Medicine Best Practices systematic review. Crit Care. 2019 Aug 9;23(1):278.

12. Welty M, Bolick BN. Eliminate Unnecessary Laboratory Work to Mitigate Iatrogenic Anemia and Reduce Cost for Patients on Extracorporeal Membrane Oxygenation. J Extra Corpor Technol. 2022 Jun;54(2):123–7.

13. Adashi EY, O’Mahony DP, Gruppuso PA. Porcine Kidney Xenotransplantation: A Rapidly Moving Frontier. The American Journal of Medicine. 2024 May;S0002934324002882.

14. Griffith Bartley P., Goerlich Corbin E., Singh Avneesh K., Rothblatt Martine, Lau Christine L., Shah Aakash, et al. Genetically Modified Porcine-to-Human Cardiac Xenotransplantation. New England Journal of Medicine. 2022 Jul 6;387(1):35–44.

15. Montgomery RA, Stern JM, Lonze BE, Tatapudi VS, Mangiola M, Wu M, et al. Results of Two Cases of Pig-to-Human Kidney Xenotransplantation. New England Journal of Medicine. 2022 May 19;386(20):1889–98.

16. Moazami N, Stern JM, Khalil K, Kim JI, Narula N, Mangiola M, et al. Pig-to-human heart xenotransplantation in two recently deceased human recipients. Nat Med. 2023 Aug;29(8):1989–97.

17. Parent B, Gelb B, Latham S, Lewis A, Kimberly LL, Caplan AL, et al. The ethics of testing and research of manufactured organs on brain-dead/recently deceased subjects. J Med Ethics. 2020 Mar;46(3):199–204.

18. Walker RL, Juengst ET, Whipple W, Davis AM. Genomic Research with the Newly Dead: A Crossroads for Ethics and Policy. J Law Med Ethics. 2014;42(2):220–31.

19. Yasko LL, Wicclair M, DeVita MA. Committee for Oversight of Research Involving the Dead (CORID): insights from the first year. Camb Q Healthc Ethics. 2004;13(4):327–37.

20. Carey RN, Jani C, Johnson C, Pearce J, Hui-Ng P, Lacson E. Chemistry Testing on Plasma Versus Serum Samples in Dialysis Patients: Clinical and Quality Improvement Implications. Clin J Am Soc Nephrol. 2016 Sep 7;11(9):1675–9.

21. Bodley T, Chan M, Clarfield L, Levi O, Longmore A, Lin W, et al. Patient Harm from Repetitive Blood Draws and Blood Waste in the ICU: A Retrospective Cohort Study. Blood. 2019 Nov 13;134(Supplement_1):57.

22. You J, Osea J, Mendoza S, Shiomi T, Gallego E, Pham B, et al. Automated and robust extraction of genomic DNA from various leftover blood samples. Analytical Biochemistry. 2023;678:115271.

23. Ostroff R, Foreman T, Keeney TR, Stratford S, Walker JJ, Zichi D. The stability of the circulating human proteome to variations in sample collection and handling procedures measured with an aptamer-based proteomics array. Journal of Proteomics. 2010 Jan 3;73(3):649–66.

24. de Almeida RW, Espírito-Santo MP, Sousa PSF, de Almeida AJ, Lampe E, Lewis-Ximenez LL. Hepatitis B virus DNA stability in plasma samples under short-term storage at 42°C. Braz J Med Biol Res. 2015 Mar 13;48(6):553–6.

25. José M, Curtu S, Gajardo R, Jorquera JI. The effect of storage at different temperatures on the stability of Hepatitis C virus RNA in plasma samples. Biologicals. 2003 Mar 1;31(1):1–8.

